# COVID-19 Vaccine Intentions in the United States—December 2020 to March 2021

**DOI:** 10.1101/2021.05.16.21257290

**Authors:** Mark É Czeisler, Shantha MW Rajaratnam, Mark E Howard, Charles A Czeisler

**Author notes:** **Corresponding Author** Mark É. Czeisler, AB, Australian-American Fulbright Scholar and PhD Candidate Turner Institute for Brain and Mental Health, Monash University Level 5, 18 Innovation Walk, Clayton Campus, Clayton, 3800, Victoria, Australia.

## Abstract

**Importance:** SARS-CoV-2 containment is estimated to require attainment of high (>80%) post-infection and post-vaccination population immunity.

**Objective:** To assess COVID-19 vaccine intentions among US adults and their children, and reasons for vaccine hesitancy among potential refusers.

**Design:** Internet-based surveys were administered cross-sectionally to US adults during December 2020 and February to March 2021 (March-2021).

**Setting:** Surveys were administered through Qualtrics using demographic quota sampling.

**Participants:** A large, demographically diverse sample of 10,444 US adults (response rate, 63.9%).

**Main Outcomes and Measures:** COVID-19 vaccine uptake, intentions, and reasons for potential refusal. Adults living with or caring for children aged 2 to 18 years were asked about their intent to have their children vaccinated. Multivariable weighted logistic regression models were used to estimate adjusted odds ratios for vaccine refusal.

**Results:** Of 5256 March-2021 respondents, 3467 (66.0%) reported they would definitely or most likely obtain a COVID-19 vaccine as soon as possible (ASAP Obtainers), and an additional 478 (9.1%) reported they were waiting for more safety and efficacy data before obtaining the vaccine. Intentions for children and willingness to receive a booster shot largely matched personal COVID-19 vaccination intentions. Vaccine refusal (ie, neither ASAP Obtainers nor waiting for more safety and efficacy data) was most strongly associated with not having obtained an influenza vaccine in 2020 (adjusted odds ratio, 4.11 [95% CI, 3.05-5.54]), less frequent mask usage (eg, rarely or never versus always or often, 3.92 [2.52-6.10]) or social gathering avoidance (eg, rarely or never versus always or often, 2.65 [1.95-3.60]), younger age (eg, aged 18-24 versus over 65 years, 3.88 [2.02-7.46]), and more conservative political ideology (eg, very conservative versus very liberal, 3.58 [2.16-5.94]); all *P*<.001.

**Conclusions and Relevance:** Three-quarters of March-2021 respondents in our large, demographically diverse sample of US adults reported they would likely obtain a COVID-19 vaccine, and 60% of adults living with or caring for children plan to have them vaccinated as soon as possible. With an estimated 27% of the US population having been infected with SARS-CoV-2, once vaccines are available to children and they have been vaccinated, combined post-infection and post-vaccination immunity will approach 80% of the US population in 2021, even without further infections.

**Key Points:** *Question:* What are COVID-19 vaccines intentions, for adults and for children under their care?

*Findings:* Two-thirds of 5256 US adults surveyed in early 2021 indicated they would obtain a COVID-19 vaccine as soon as possible. Intentions for children and booster vaccines largely matched personal vaccine intentions. Refusal was more common among adults who were younger, female, Black, very politically conservative, less educated, less adherent with COVID-19 prevention behaviors (eg, wearing masks), had more medical mistrust, or had not received influenza vaccines in 2020.

*Meaning:* Tailored vaccine promotion efforts and vaccine programs may improve vaccine uptake and contribute to US immunity against COVID-19.

## Introduction

As of mid-May 2021, the US Food & Drug Administration (FDA) has granted Emergency Use Authorization for 3 coronavirus disease 2019 (COVID-19) vaccines (Pfizer-BioNTech, Moderna, and Janssen),^1^ and nearly 60% of US adults have been fully (120 million) or partially (35 million) vaccinated.^2^ Consistent with vaccine prioritization,^3^ ∼85% of adults aged 65-plus years have received vaccines.^2^ Early indicators demonstrate high efficacy of these vaccines in reducing severe acute respiratory coronavirus syndrome 2 (SARS-CoV-2) transmission and severe COVID-19 outcomes.^4-14^ These represent remarkable public health and scientific achievements. Yet, several obstacles remain to containing COVID-19 in the US and globally.

First, until recently, the rate-limiting steps in the US have been vaccine supplies and delivery capacity. Only now that supply is plentiful has vaccine hesitancy started to present as a barrier to vaccination *en masse*. Prior studies provided information for tailored educational programs to enhance informed COVID-19 vaccine decision-making.^15-23^ Understanding groups that remain disproportionately vaccine hesitant, and common reasons for hesitancy, are critical to promote vaccination.

Second, initial COVID-19 vaccine clinical trials excluded individuals aged under 16 years and pregnant persons. Encouragingly, recent studies have led to Emergency Use Authorization of the Pfizer-BioNTech COVID-19 vaccine for children aged 12-15 years,^24,25^ with studies of additional age groups underway. Moreover, COVID-19 mRNA vaccines are safe and effective in pregnant people^26,27^ and confer immunity to neonates,^28^ which is particularly important given substantially elevated risk of adverse maternal and neonatal health outcomes from SARS-CoV-2 infection.^29-33^

Third, vaccine-evasive coronavirus variants could threaten post-vaccination immunity. Fortunately, the developed vaccines are effective against most common variants,^10,34^ though more evasive variants have started to appear,^35^ some of which may require modified COVID-19 vaccine boosters.^36,37^ Understanding COVID-19 booster vaccine intentions is therefore important.

We therefore assessed COVID-19 vaccine uptake, intentions, and reasons for hesitancy in a large, diverse sample of US adults, including pregnant people. We examined child vaccine intentions among parents and caregivers, and willingness to receive variant-protective COVID-19 booster vaccines.

## Methods

### Setting and Participants

From December 6-27, 2020 (December-2020) and February 16 to March 8, 2021 (March-2021), anonymous, Internet-based surveys were administered to non-overlapping 18-plus year-old US residents for The COVID-19 Outbreak Public Evaluation (COPE) Initiative (www.thecopeinitiative.org). Surveys were administered to panels maintained by Qualtrics. Nonprobability demographic quota sampling and survey weighting were employed to match national US adult population 2019 American Community Survey estimates for sex, age, and race/ethnicity. Weighted values are reported unless specified.

### Survey Instrument

The December-2020 and March-2021 survey instruments comprised 136 and 160 items, respectively, and included questions about demographics, pandemic-related attitudes and behaviors, and mental health. Respondents were not informed of survey topics prior to commencement.

### Key Definitions

#### Vaccine Intentions

COVID-19 vaccination intentions were assessed using the question, “If an FDA-approved vaccine to protect against COVID-19 were widely accessible, would you get one as soon as possible?” Respondents answered using a five-item Likert scale: “No, definitely not,” “Unlikely,” “Maybe/Not sure,” “Most likely,” or “Yes, definitely”. March-2021 respondents could also answer that they had been vaccinated against COVID-19. Respondents who selected “No, definitely not,” “Unlikely,” or “Maybe/Not sure” selected among 8 reasons for not obtaining a vaccine as soon as possible (ASAP), with multiple selections allowed: waiting for more safety and efficacy data, low COVID-19 risk perception, beliefs the vaccine would not protect against COVID-19, the approval process was rushed, or that all vaccines are dangerous, concern of a hidden purpose, religious refusal, and other. March-2021 respondents who reported living with or caring for persons aged 2 to 18 years were asked about COVID-19 vaccination intentions for their children. All March-2021 respondents were asked about potential COVID-19 booster intentions.

#### Characteristics

Demographic characteristics assessed included sex, age, race/ethnicity, education attainment, pregnancy, parental or unpaid caregiver roles, and political ideology. Medical mistrust was assessed using the Medical Mistrust Index (MMI),^38^ with responses categorized into 4 levels (0-6, 7-13, 14-17, and 18-21). Higher scores reflect more mistrust. Respondents reported whether they had received an influenza vaccine last year or ever tested SARS-CoV-2-positive, and past-week frequency of mask usage in public and avoidance of 10-plus-person gatherings using 5-item Likert scales: never, rarely, sometimes, often, and always.^39^

## Statistical Analysis

Intentions to receive COVID-19 vaccines in December-2020 and March-2021 were grouped as Decliners (“No, definitely not” or “Unlikely”), Undecideds (“Maybe/Not sure”), or ASAP Obtainers (“Most likely” or “Yes, definitely,” plus March-2021 respondents who had already been vaccinated). A category of Overall Obtainers was created as ASAP Obtainers, plus respondents waiting for more safety and efficacy data (a subset of Decliners and Undecideds). Chi-square tests with design effect correction factors were used to test for differences between March-2021 subgroups (eg, male versus female respondents), and between the December-2020 and March-2021 samples within subgroups (eg, non-overlapping female respondents over time). Bonferroni adjustments of 9 and 33, respectively, were applied to account for multiple comparisons.

Weighted logistic regression models were used to estimate unadjusted and adjusted odds ratios (aORs) and 95% confidence intervals (CIs) for vaccine Refusal (ie, Decliners, minus those waiting for safety and efficacy data) among March-2021 respondents. Multivariable models included sex, age group, race/ethnicity, education attainment, parental or unpaid caregiving roles, political ideology, and health insurance as covariates. To avoid collinearity, separate models were run for frequency of mask usage and avoiding gatherings, MMI score, and past-year influenza vaccination. Among female respondents of childbearing age, a regression was run based on pregnancy status. To account for 11 comparisons, point estimates are reported with 95% CIs that were estimated at the 99.545% confidence level and Bonferroni-adjusted (n=11) *P*.

Among vaccine Decliners and Undecideds, crosstabs of select characteristics and reasons for hesitancy were calculated. Intentions for vaccinating children among March-2021 respondents living with or caring for children, and acceptance of potential vaccine booster doses among all March-2021 respondents, were described based on personal vaccine intentions. To identify factors associated with indecision versus complete Refusal, weighted logistic regression models were used to estimate ORs and aORs for indecision (ie, responding Maybe versus Unlikely or Definitely not, or selecting that they were waiting for more safety and efficacy data versus other reasons). Multivariable models included all demographics listed in the primary regression models. To account for 7 comparisons, point estimates are reported with 95% CIs that were estimated at the 99.286% confidence level and Bonferroni-adjusted (n=7) *P*.

Data were cleaned in Python version 3.7.8 (Python Software Foundation). Calculations were made in R version 4.0.2 (The R Project for Statistical Computing) using the R survey package version 3.29. Statistical significance was set at 2-sided *P*<.05. Detailed methods are in the Supplement.

### Study Review

Respondents provided informed electronic consent. The Monash University Human Research Ethics Committee (Melbourne, Australia) reviewed and approved the protocol. Given exclusive recruitment of US residents in 2021, the Mass General Brigham Institutional Review Board (Boston, Massachusetts) also reviewed the protocol prior to the March-2021 wave and determined that this public health surveillance activity did not require institutional review board review. This study followed the American Association for Public Opinion Research guidelines.

## Results

Overall, 10,469 of 16,384 (response rate, 63.9%) invited eligible adults completed surveys. Of these, 10,444 (99.8%) reported sex, age, race, and ethnicity used for survey weighting and were included in this analysis (eFigure 1). Of analyzed respondents, 5188 completed December-2020 surveys, and 5256 completed March-2021 surveys (Table 1).

**Table 1.**
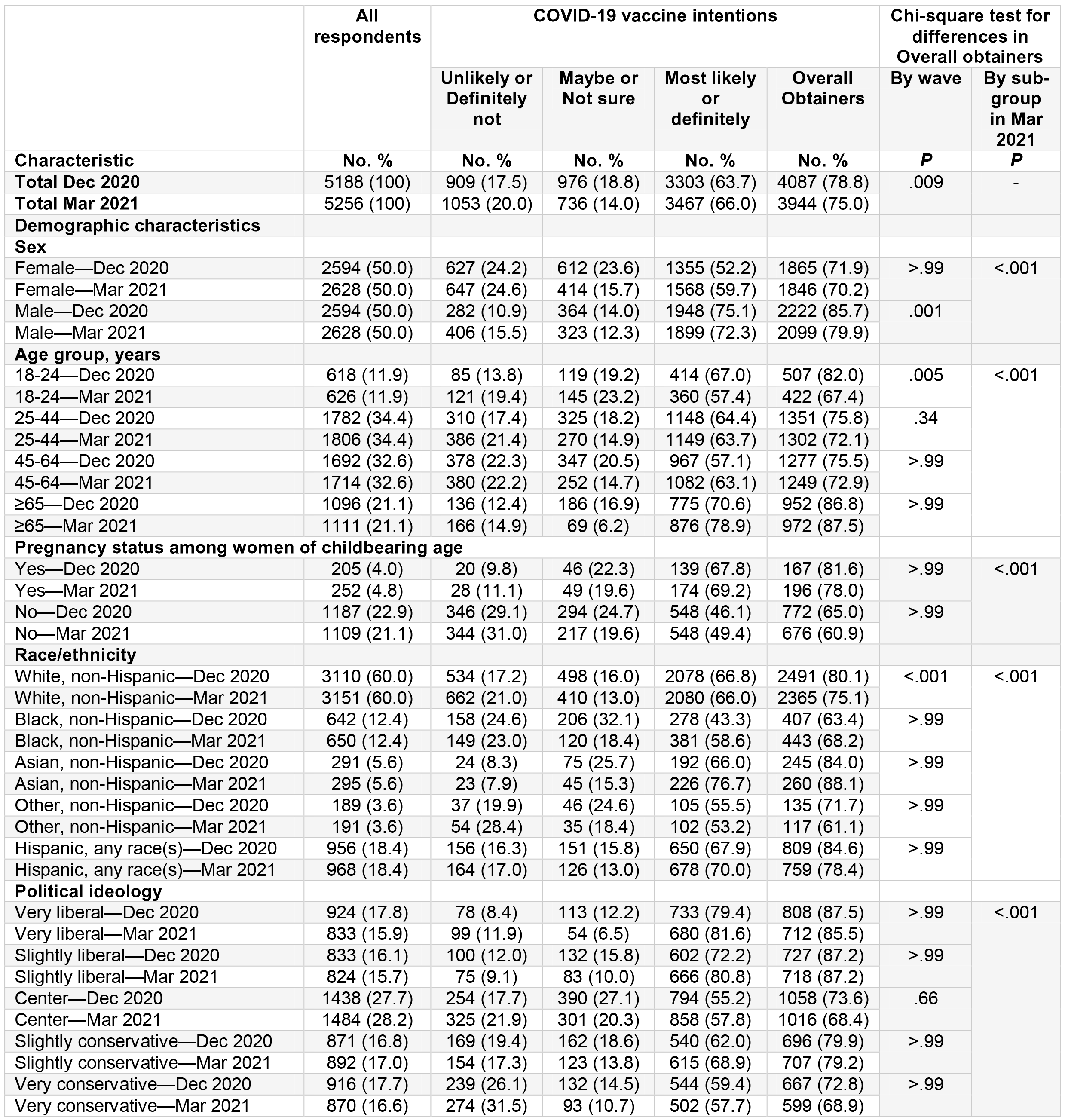
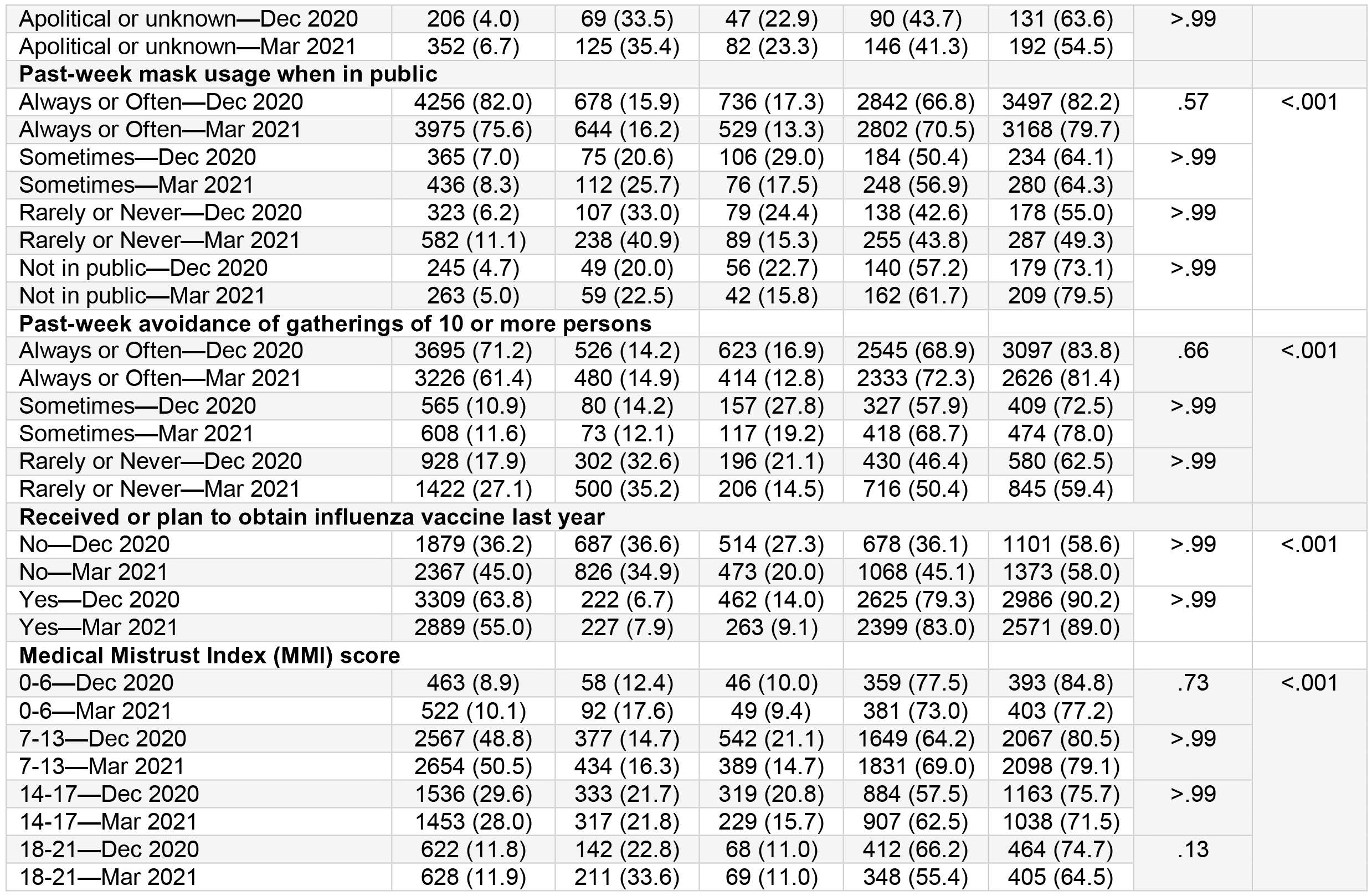
COVID-19 vaccine intentions among US adults—December 2020 and March 2021.

Of December-2020 respondents, 909 (17.5%) respondents were Decliners, 976 (18.8%) were Undecideds, and 3303 (63.7%) were ASAP Obtainers (Table 1). Of 5256 March-2021 respondents, 1053 (20.0%) were Decliners, 736 (14.0%) were Undecideds, and 3467 (66.0%) were ASAP Obtainers. The percentage of Overall Obtainers was lower in March-2021 as compared to December-2020 (3944 of 5256 [75.0%], 4087 of 5188 [78.8%], respectively, *P*=.009). Within demographic subgroups across waves, the prevalence of Overall Obtainers was lower in March-2021 as compared to December-2020 among male respondents (2099 of 2628 [79.9%], 2222 of 2594 [85.7%], *P*=.001), adults aged 18 to 24 years (422 of 626 [67.4%], 507 of 618 [82.0%], *P*=.005), and White respondents (2365 of 3151 [75.1%], 2491 of 3110 [80.1%], *P*<.001). Between all subgroups in Table 1, the prevalence of Overall Obtainers differed significantly. In general, the prevalence was higher among respondents who were male versus female, older versus younger, Asian or Hispanic compared with Black, liberal versus conservative, and, among female respondents of childbearing age, those who were pregnant versus those who were not. The prevalence of Overall Obtainers was also higher among respondents who wore masks in public or avoided social gatherings more frequently, had received or planned to receive the influenza vaccine, and had lower levels of medical mistrust.

Multivariable analysis of March-2021 respondents revealed that odds of vaccine Refusal were highest among adults who had not received an influenza vaccine (aOR, 4.11 [95% CI, 3.05-5.54], *P*<.001) (Figure 1, eTable 1). Refusal was also positively associated with less frequent mask usage (eg, rarely or never versus always or often, .92 [2.52-6.10], *P*<.001) or gathering avoidance (eg, rarely or never versus always or often, 2.65 [1.95-3.60], *P*<.001), younger age (eg, aged 18-24 versus 65-plus years, 3.88 [2.02-7.46], *P*<.001), more conservative political ideology (eg, very conservative versus very liberal, 3.58 [2.16-5.94], *P*<.001), lower education attainment (eg, high school diploma or less versus more than bachelor’s degree, 3.43 [2.11-5.59], *P*<.001), higher levels of medical mistrust (MMI scores 18-21 versus 0-6, 2.11 [1.10-4.07], *P*<.001), female versus sex (1.51 [1.16-1.96], *P*<.001), and Black (1.60 [1.10-2.33], *P*=.004) or other (1.99 [1.15-3.42], *P*=.004) versus White race/ethnicity. Conversely, lower odds of vaccine refusal were observed for respondents who were of Asian versus White race/ethnicity (.42 [.20-.90], *P*=.013), and among multigenerational caregivers versus non-caregivers (.51 [.35-.74], *P*<.001). Unadjusted ORs are in eTable 2.

**Figure 1.**
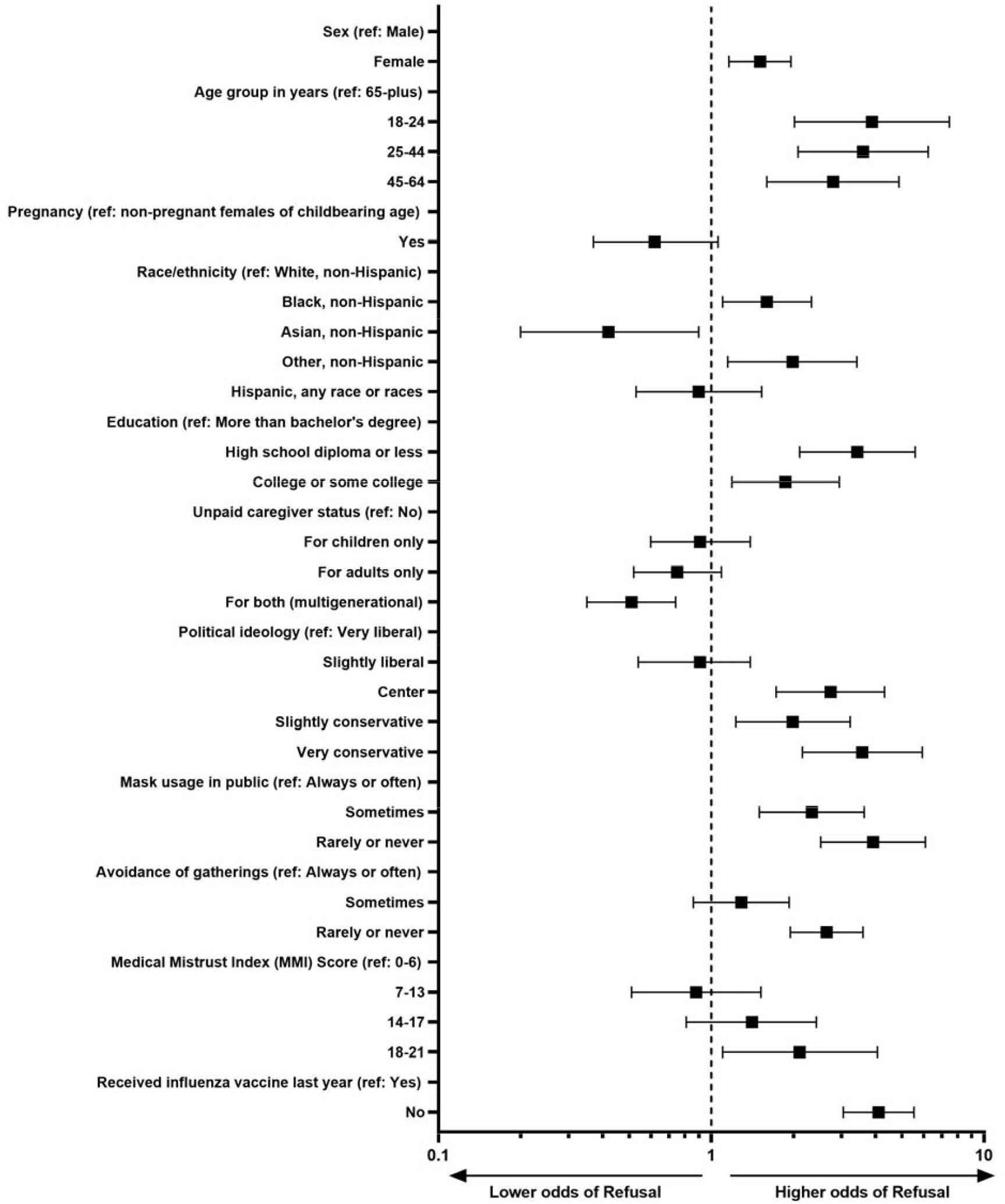
Adjusted odds ratios for COVID-19 vaccine refusal among US adults—March 2021.

Among 1789 March-2021 Undecideds or Decliners, common reasons for potentially not being ASAP Obtainers were concern that the vaccine may be risky due to rushed approval (41.8%), plans to wait 6-12 months for safety and efficacy data (26.7%), concern of a hidden purpose (25.0%), and belief that the vaccine would not offer protection from COVID-19 (24.3%) or low COVID-19 risk perception (18.0%) (Table 2). Comparing March-2021 (n=1789) versus December-2021 (n=1885) respondents who were Undecideds or Decliners, the percentage who were planning to wait for more data decreased over time (478 [26.7%], 784 [41.6%], respectively, *P*<.001), as did the percentage who reported concerns that the approval process had been rushed (746 [41.7%], 919 [48.7%], *P*=.004). The percentage who were concerned of a hidden purpose was increased over time (446 [24.9%], 358 [19.0%], *P*=.008).

**Table 2.**
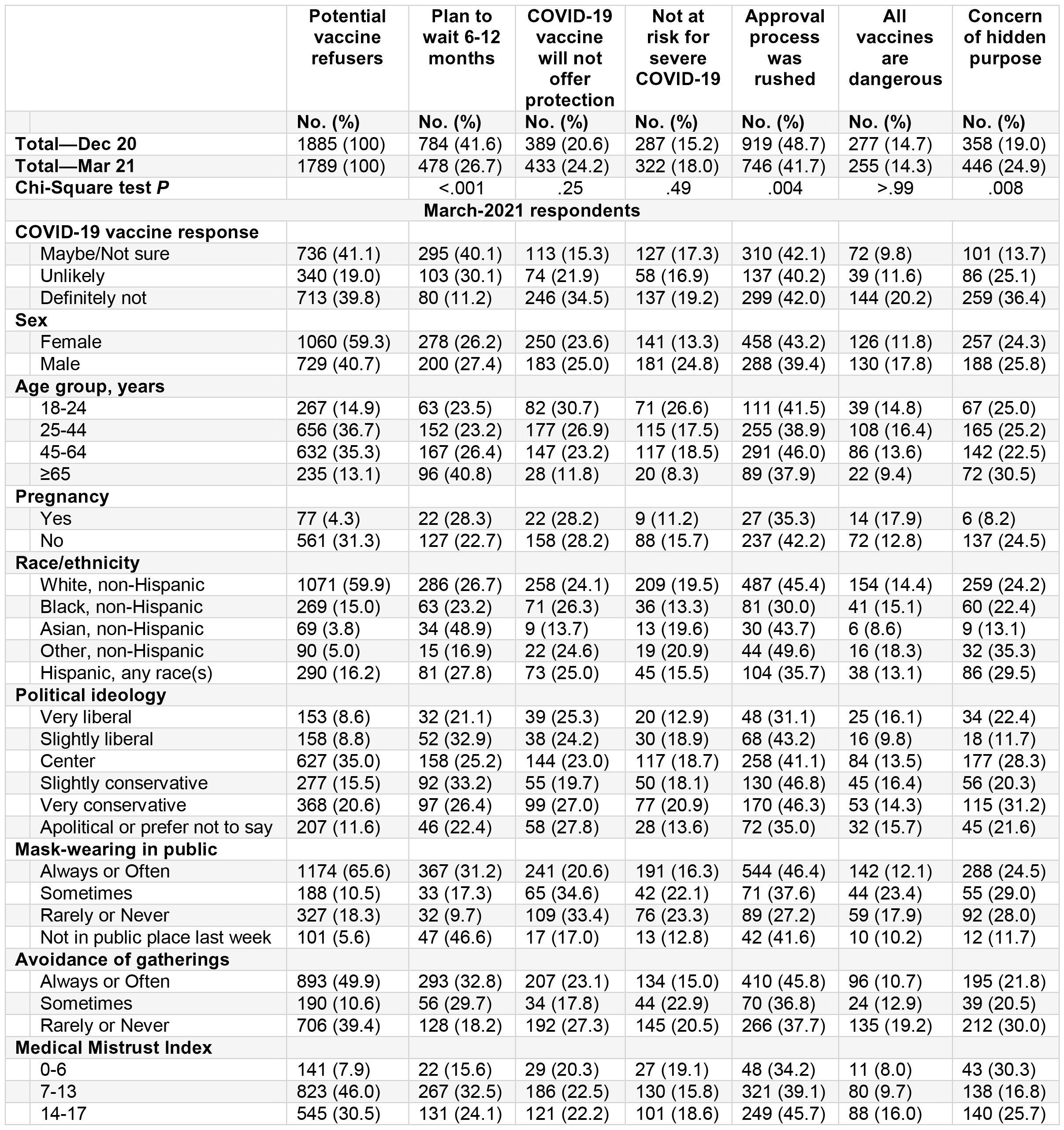
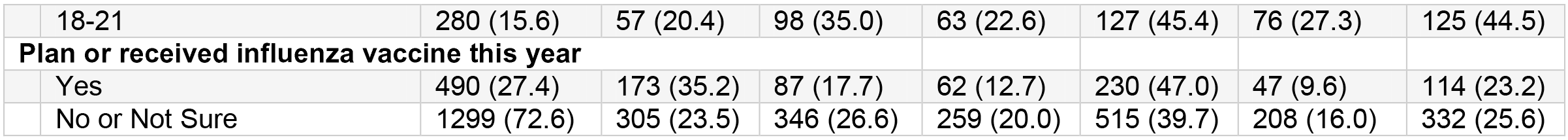
Reasons for potential COVID-19 vaccine refusal among US adults—December 2020 and March 2021.

Of March-2021 Undecideds or Decliners, aORs for being an Undecided rather than a Decliner were higher among individuals aged 18-24 versus 65-plus years (aOR, 2.30 [95% CI, 1.08-4.90], *P*=.021), multigenerational caregivers versus non-caregivers (1.58 [1.01-2.47], *P*=.042), and those with more centrist versus very conservative political ideology (Figure 2A, eTable 3). Lower aORs for being an Undecided were found for individuals with a high school diploma or less versus more than a bachelor’s degree (.42 [.22-.81], *P*=.003). Regarding waiting for more safety and efficacy data, aORs were significantly lower for adults aged 25-44 versus 65-plus years (.41 [.19-.89], *P*=.013) (Figure 2B, eTable 5). No other significant demographic associations were found. Unadjusted ORs are in eTables 4 and 6.

**Figure 2.**
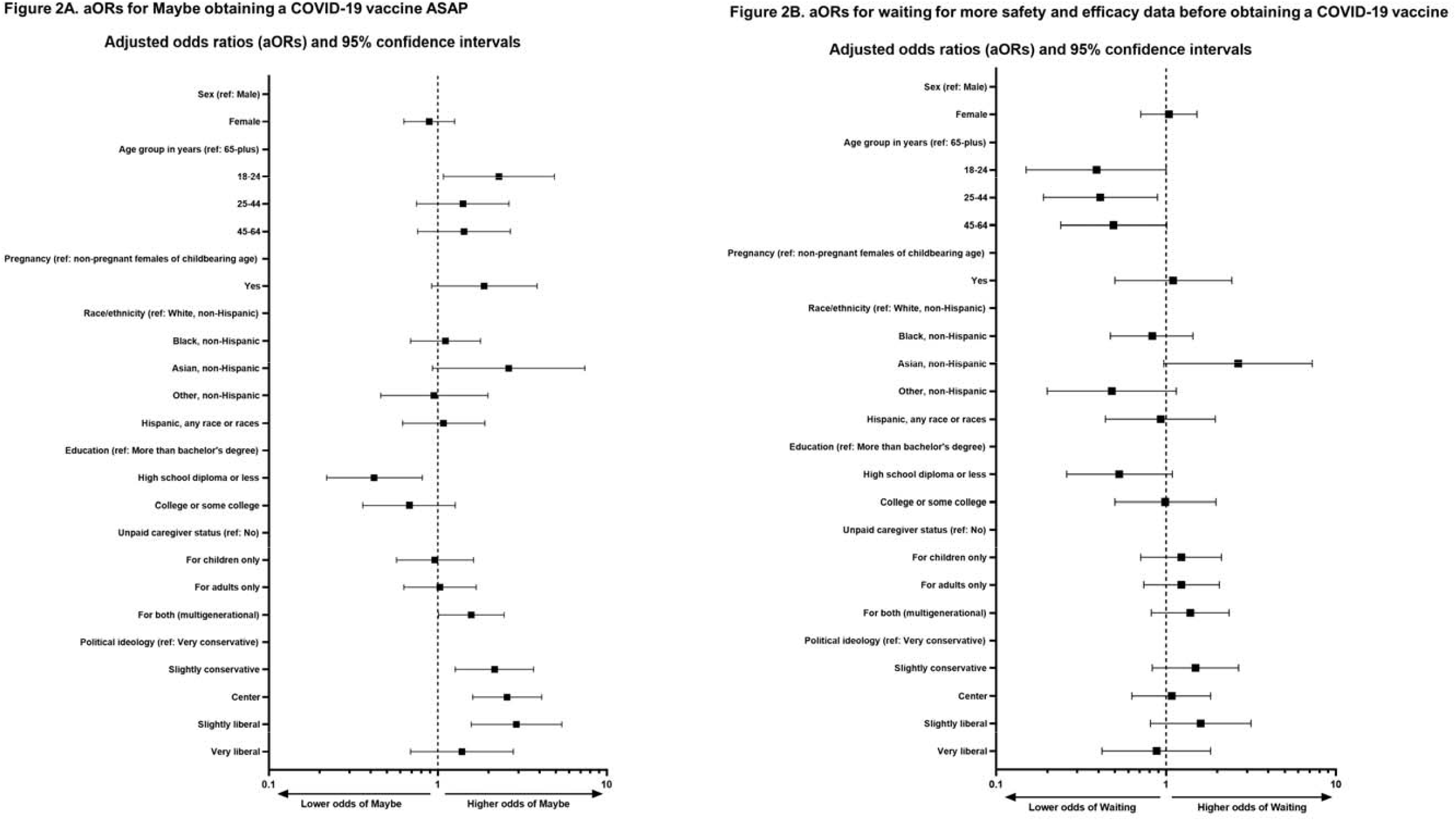
Adjusted odds ratios for responding Maybe or waiting for more safety and efficacy data before obtaining a COVID-19 vaccine among US adult vaccine Refusers —March 2021.

Among 2160 March-2021 respondents living with or caring for children aged 2-18 years, intentions to vaccinate those children were similar to those for adults (1305 [60.4%] ASAP Obtainers 463 [18.1%] Undecideds, 463 [21.4%] Decliners) (Figure 3A). Of 1305 ASAP Obtainers for their children aged 2-18 years, 1221 (93.5%) were ASAP Obtainers for themselves, while only 39 (3.0%) were Decliners for themselves. Conversely, of 463 Decliners for their children aged 2-18 years, only 119 (25.7%) were ASAP Obtainers for themselves, while 261 (56.5%) were Decliners for themselves. Similar relationships with personal vaccine intentions were found for booster vaccine intentions. Of 3074 March-2021 ASAP booster Obtainers, 2928 (95.2%) were ASAP Obtainers of the original COVID-19 vaccine, while Decliners accounted for just 49 (1.6%) of these ASAP booster Obtainers (Figure 3B).

**Figure 3.**
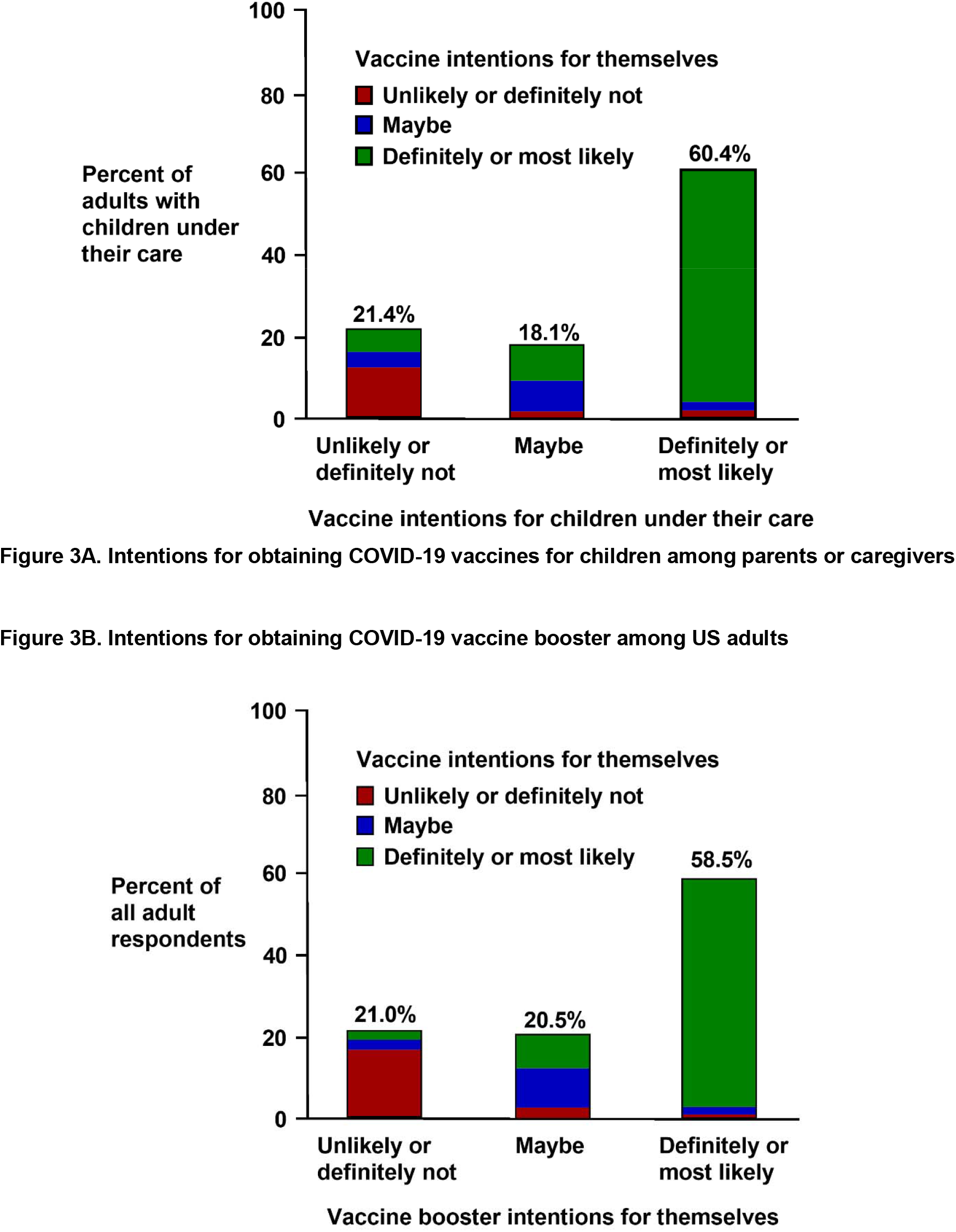
Intentions for obtaining COVID-19 vaccines for children and potential vaccine boosters to protect against variants—March 2021.

## Discussion

Nearly two-thirds of 5256 US adults surveyed during mid-February to early March 2021 reported they had obtained or would definitely or most likely obtain an FDA-approved COVID-19 vaccine as soon as possible, with up to three-quarters likely obtainers when including individuals waiting for more safety and efficacy data. Given that approximately 60% of the US adult population has received at least one dose of the COVID-19 vaccine as of May 15, 2021,^2^ these data suggest that less than one-quarter of the remaining unvaccinated US adults planned to obtain vaccines ASAP, and less than one-half planned to ever be vaccinated against COVID-19. Vaccine Refusal was highest among adults who were younger, female, Black or other (versus White) race/ethnicity, very conservative politically, those with lower education attainment, more medical mistrust, lesser COVID-19 prevention behavior adherence, and those who had not received influenza vaccines. As the US vaccine rollout faces the barriers of vaccine hesitancy in the majority of the remaining unvaccinated US adults, vaccine promotion activities tailored for these groups may improve uptake.

Nearly 70% of March-2021 pregnant females of childbearing age were ASAP obtainers. Early COVID-19 vaccine safety and efficacy among pregnant persons are encouraging.^28,29^ Higher risk of adverse clinical outcomes among pregnant persons and their neonates among individuals with SARS-CoV-2 infection,^29,30,33^ including a 22-fold increased risk of maternal mortality,^40^ underscores the importance of ensuring vaccination access for this willing, at-risk population. Parental decisions about obtaining COVID-19 vaccines for their children largely matched their personal intentions, revealing that groups identified as potential vaccine refusers will likely do the same for their children. Our findings indicate that parents and caregivers intend to use the vaccine distribution infrastructure to vaccinate children under their care once emergency use authorizations are revised to expand the range of approved ages. This is particularly encouraging given that young adults and children facilitate SARS-CoV-2 transmission^41-44^ and have sustained regional outbreaks.^45-48^

According to Sanche *et al*. in the CDC’s *Emerging Infectious Diseases*, with an estimated R_0_ of 5.7, SARS-CoV-2 containment requires approximately 82% of the population to obtain post-vaccination or post-infection immunity.^49^ In Israel, which was among the world leaders in vaccination rate through mid-May 2021, COVID-19 deaths declined from 70 deaths per day in January 2021 to 0.^50^ New SARS-CoV-2 infections and COVID-19 deaths have also dropped considerably in the US, where more than half of US adults have received at least one COVID-19 vaccine dose. Given that the CDC estimates that at least 27% of US adults had been infected with SARS-CoV-2 as of December 2020,^51,52^ and considerably more since then, even if only 33% of these were vaccine refusers, combined post-vaccination and post-infection immunity among US adults should approach 76%. Thus, if half of the 9% of individuals waiting for more safety and efficacy data were to obtain the vaccine, approximately 80% of US adults would have some SARS-CoV-2 immunity. Moreover, as SARS-CoV-2 infection is particularly prevalent among those who were non-adherent with CDC COVID-19 prevention guidance,^53,54^ this group of mostly vaccine Refusers likely has more post-infection immunity. Achieving population-level immunity, however, depends on vaccination or infection of children. Fortunately, 60% of surveyed parents or caregivers for children reported being ASAP Obtainers for their children.

To achieve high levels of immunity, engaging the Undecideds (15% of March-2021 respondents) will be critical. Young age, more centrist political ideology, and multigenerational caregiver status were associated with being Undecided, and may represent high-yield demographics to incentivize uptake. Interestingly, only one-quarter of young adult Undecideds indicated that they were waiting for more vaccine safety and efficacy data, suggesting that alternative incentives should be reviewed based on prior immunization programs^55-59^ and investigated during the current rollout^60,61^ (eg, monetary incentives,^62^ vaccine mandates for return to campus, employer or workplace vaccination programs, or easing restrictions for vaccinated persons, such as those reported in recent CDC guidance^63^ and the European Union’s international travel ban for those fully vaccinated^64^). Concurrently, ensuring equitable access to vaccines may reduce disparities—particularly regarding internet connectivity and technology usability and literacy.^65^

Monitoring and responding to SARS-CoV-2 variants will be essential. Development of vaccine boosters to combat vaccine-evasive variants is underway. Our results suggest that acceptance of COVID-19 vaccine boosters will largely reflect overall COVID-19 vaccination trends. To avoid further COVID-19 health disparities, improving vaccine uptake among groups with high levels of vaccine refusal will prove important. Furthermore, the race against variants will occur globally.^66,67^ The US is among the high-income countries with abundant vaccine supply, while many low- and middle-income countries have struggled to initiate vaccination campaigns.^68,69^

Strengths of this study include assessment of COVID-19 vaccine and booster intentions in large, demographically diverse samples of US adults at multiple timepoints, and inclusion of diverse characteristics. Limitations include self-reported metrics that may not correlate with future behavior and Internet-based survey methods that may not fully represent the US population. However, our data for the prevalence of COVID-19 vaccine recipients as of mid-February to early March 2021 were consistent with nationwide surveillance data,^70^ and 88.7% of respondents who had received 1 dose in a 2-dose regimen indicated that they planned to complete the series, consistent with CDC surveillance data (88.0%).^71^

Projections of US population immunity are contingent on assumptions.^72,73^ First, post-vaccination population immunity requires efficacy against infection above 80%,^74^ well below current estimates.^6^ Second, evidence from other coronaviruses^75,76^ and preliminary reports of SARS-CoV-2 re-infection^77-79^ or breakthrough infections among fully vaccinated individuals^80-83^ suggest both vaccination- and infection-derived immunity may be transient, requiring re-vaccination. Third, current FDA-approved vaccines are not authorized for children aged under 12 years. Fourth, most current vaccines require multiple doses for maximal efficacy, presenting a barrier to distribution.^84^ However, nearly 90% of people in 2-dose COVID-19 vaccine regiments received both doses, and more than 95% of completers did so within the recommended interval between the first and second doses.^71^ Finally, considerable regional differences in vaccination rates will affect local transmission of the SARS-CoV-2 viral infections.

Our findings reveal that vaccine hesitancy is unlikely to prevent the US from achieving high levels of immunity against COVID-19 in 2021, and that intentions for vaccination of children and obtaining boosters largely match personal vaccine intentions. Vaccine education campaigns tailored for Undecideds, coupled with robust vaccine distribution programs, could enhance vaccine obtainment and assist in controlling the COVID-19 pandemic in the US.

## Data Availability

De-identified participant data can be made available by contacting the corresponding author upon reasonable request. Reuse is only permitted following a written agreement from the corresponding author and primary Institution.

## Author Contributions

Mr Czeisler had full access to all of the data in the study and takes responsibility for the integrity of the data and the accuracy of the data analysis.

*Concept and design:* All authors.

*Acquisition, analysis, or interpretation of data:* All authors.

*Drafting of the manuscript:* Czeisler, Czeisler.

*Critical revision of the manuscript for important intellectual content:* All authors.

*Statistical analysis:* Mr Czeisler.

*Obtained funding:* All authors.

*Supervision:* Howard, Rajaratnam.

## Conflict of Interest Disclosures

Mr Czeisler reported personal fees from Vanda Pharmaceuticals Inc. Dr Rajaratnam reported receiving grants and personal fees from Cooperative Research Centre for Alertness, Safety and Productivity, receiving grants and institutional consultancy fees from Teva Pharma Australia, and institutional consultancy fees from Vanda Pharmaceuticals, Circadian Therapeutics, BHP Billiton, and Herbert Smith Freehills. Dr Czeisler reported receiving grants and personal fees from Teva Pharma Australia, receiving grants from the National Institute of Occupational Safety and Health R01-OH-011773, receiving personal fees from and equity interest in Vanda Pharmaceuticals Inc, educational and research support from Philips Respironics Inc, an endowed professorship provided to Harvard Medical School from Cephalon, Inc, an institutional gift from Alexandra Drane, and a patent on Actiwatch-2 and Actiwatch-Spectrum devices with royalties paid from Philips Respironics Inc. Dr Czeisler’s interests were reviewed and managed by Brigham and Women’s Hospital and Partners HealthCare in accordance with their conflict of interest policies. Dr Czeisler also served as a voluntary board member for the Institute for Experimental Psychiatry Research Foundation, Inc. No other disclosures were reported.

## Funding/Support

Funding for survey data collection was supported in part by research grants from the CDC Foundation (Atlanta, Georgia) with funding from BNY Mellon (New York, New York), and from WHOOP, Inc (Boston, Massachusetts), and Hopelab, Inc (San Francisco, California) to Monash University acting through its Faculty of Medicine, Nursing and Health Sciences, and by institutional support from Philips Respironics Inc to Brigham & Women’s Hospital, the Turner Institute for Brain and Mental Health, Monash University, and Institute for Breathing and Sleep, Austin Hospital. Mr Czeisler gratefully acknowledges support from a 2020 Australian-American Fulbright Scholarship funded by The Kinghorn Foundation. Dr Czeisler is the incumbent of an endowed professorship provided to Harvard University by Cephalon, Inc.

### Role of the Funder/Sponsor

The funders had no role in the design and conduct of the study; collection, management, analysis, and interpretation of the data; preparation or approval of the manuscript; and decision to submit the manuscript for publication.

## Notes

### Author Declarations

Respondents provided informed electronic consent. The Monash University Human Research Ethics Committee (Melbourne, Australia) reviewed and approved the protocol. Given exclusive recruitment of US residents in 2021, the Mass General Brigham Institutional Review Board (Boston, Massachusetts) also reviewed the protocol prior to the March-2021 wave and determined that this public health surveillance activity did not require institutional review board review.

