## Supplement for "COVID-19 Vaccine Intentions in the United States—December 2020 to March 2021"

**Title**

**Contents**

**eAppendix. Supplementary Methods**

**eFigure. eFigure 1. Flow of Survey Respondents—December 2020 and March 2021**

**eTable 1. Adjusted odds ratios for COVID-19 vaccine refusal among US adults—March 2021**

**eTable 2. Unadjusted odds ratios for COVID-19 vaccine refusal among US adults—March 2021**

**eTable 3. Adjusted odds ratios for responding Maybe to obtaining a COVID-19 vaccine as soon as possible among US adult vaccine Refusers—March 2021**

**eTable 4. Unadjusted odds ratios for responding Maybe to obtaining a COVID-19 vaccine as soon as possible among US adult vaccine Refusers —March 2021**

**eTable 5. Adjusted odds ratios for waiting for more safety and efficacy data before obtaining a COVID-19 vaccine among US adult vaccine Refusers —March 2021**

**eTable 6. Unadjusted odds ratios for waiting for more safety and efficacy data before obtaining a COVID-19 vaccine among US adult vaccine Refusers —March 2021**

**eAppendix. Supplementary Methods**

### Recruitment Methodologies

Qualtrics recruitment methodologies include digital advertisements and promotions, word of mouth, and membership referrals, social networks, television and radio advertisements, and offline, mail-based approaches. Potential respondents received invitations and could opt to participate by activating a survey link directing them to the participation information and consent page preceding the survey. Ineligible respondents who did not meet inclusion criteria (eg, aged below 18 years, or exceeded pre-specified demographic quotas) were disempaneled from the survey.

### Demographic Categories

Sex was analyzed as male or female. Age was assessed as a numerical variable and categorized as ages 18 to 24 years, 25 to 44 years, 45 to 64 years, or 65 years or over. Race and ethnicity were assessed separately and combined for analysis into 5 mutually exclusive groups: non-Hispanic White (White), non-Hispanic Black (Black), Hispanic or Latino (Hispanic), non-Hispanic Asian (Asian), and non-Hispanic Other race (including multiple races). Education attainment was assessed using the 2010 Census question and combined for analysis into high school diploma or less, college or some college, and more than bachelor’s degree. Pregnancy was assessed among female respondents who were of childbearing age (less than 49 years). Unpaid caregiver (caregiver) status was categorized as non-caregivers or caregivers for adults only, children only, or both (multigenerational). Political ideology was assessed as very liberal, slightly liberal, neither liberal nor conservative (center), slightly conservative, very conservative, and prefer not to answer (unknown).

**Reporting Race/Ethnicity**

Race and ethnicity were assessed among survey respondents with separate questions and options defined by the investigators based on US Census classifications. The race and ethnicity questions follow.

***1.*** *What is your race? (Select all that apply)*
   ***a.*** *American Indian or Alaskan Native*
   ***b.*** *Asian*
   ***c.*** *Black or African American*
   ***d.*** *Native Hawaiian or other Pacific Islander*
   ***e.*** *White*
   ***f.*** *Other*

*Please use the categories that most reflect your recognition in the community for purposes of reporting mixed racial and/or ethnic origins.*

*American Indian or Alaskan Native: a person having origins in any of the original peoples of North, Central, or South America, and maintains tribal affiliations or community attachment.*

*Asian: A person having origins in any of the original peoples of the Far East, Southeast Asia, or the Indian subcontinent including, for example, Cambodia, China, India, Japan, Korea, Malaysia, Pakistan, the Philippine Islands, Thailand, and Vietnam.*

*Black or African American: A person having origins in any of the black racial groups of Africa.*

*Native Hawaiian or Pacific Islander: A person having origins in any of the original peoples of Hawaii, Guam, Samoa, or other Pacific Islands.*

*White: A person having origins in any of the original peoples of Europe, North Africa, or the Middle East.*

***2.*** *What is your ethnicity? (Select one)*
   ***a.*** *Hispanic or Latino*
   ***b.*** *Not Hispanic or Latino*

*Hispanic or Latino: A person of Cuban, Mexican, Puerto Rican, South or Central American, or other Spanish culture or origin regardless of race.*

For this analysis, race and ethnicity were combined into the following categories: White, non-Hispanic; Black, non-Hispanic; Asian, non-Hispanic; Multiple races or Other race, non-Hispanic; and Hispanic, any race or races.

### eFigure 1. Flow of Survey Respondents—December 2020 and March 2021

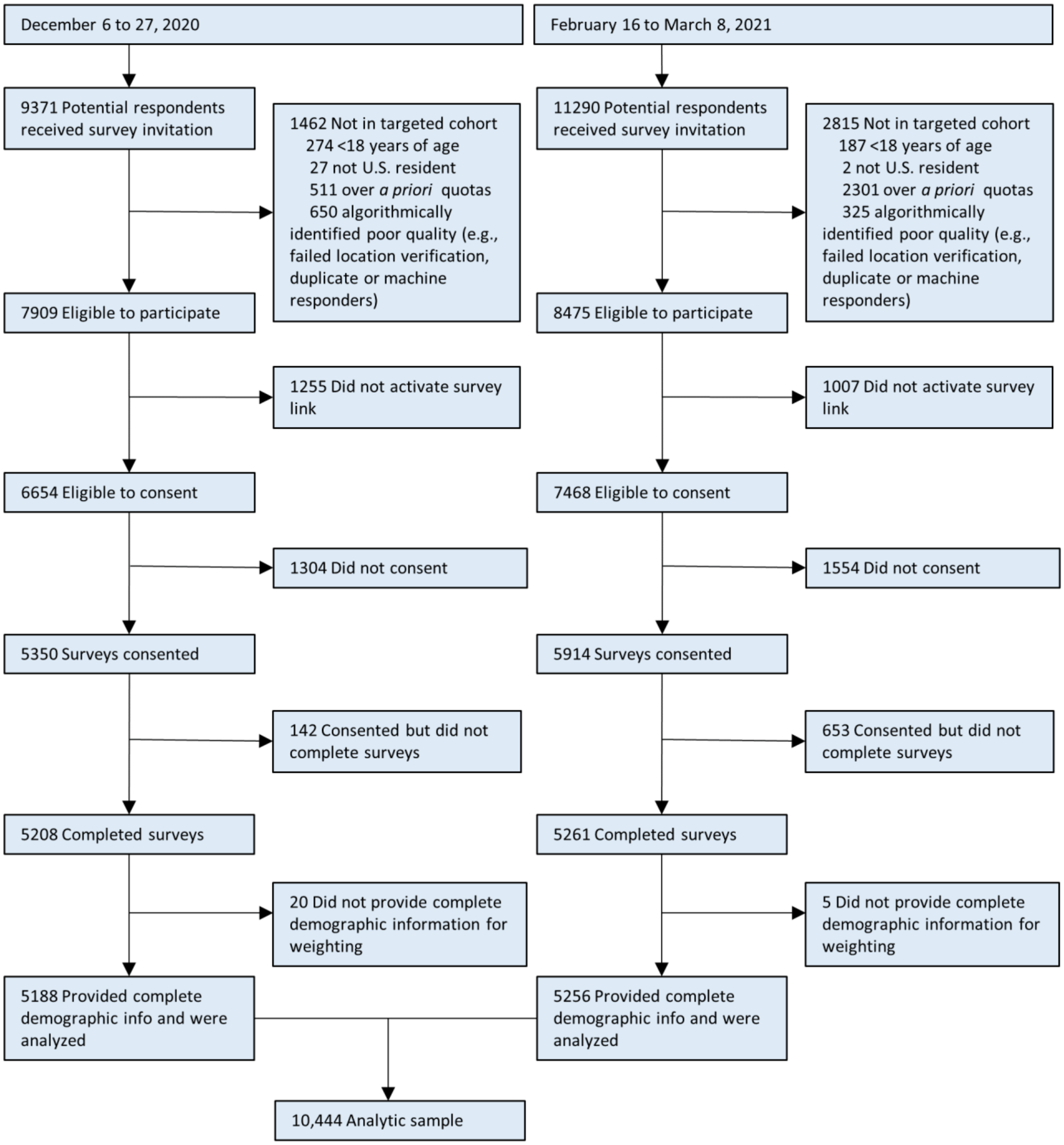

### eTable 1. Adjusted odds ratios for COVID-19 vaccine refusal among US adults—March 2021

| | | Vaccine Refusal | |
|---|---|---|---|
| | | aOR (95% CI) | *P* |
| **Sex (reference: Male)** | | | |
| | Female | 1.51 (1.16, 1.96) | <.001 |
| **Age group, years (reference: ≥65)** | | | |
| | 18-24 | 3.88 (2.02, 7.46) | <.001 |
| | 25-44 | 3.60 (2.08, 6.24) | <.001 |
| | 45-64 | 2.80 (1.60, 4.88) | <.001 |
| **Pregnancy (reference: non-pregnant females aged 18-49 years)** | | | |
| | Yes | .62 (.37, 1.06) | .14 |
| **Race/ethnicity (reference: White, non-Hispanic)** | | | |
| | Black, non-Hispanic | 1.60 (1.10, 2.33) | .004 |
| | Asian, non-Hispanic | .42 (.20, .90) | .013 |
| | Other, non-Hispanic | 1.99 (1.15, 3.42) | .004 |
| | Hispanic, any race or races | .90 (.53, 1.53) | >.99 |
| **Education (reference: More than Bachelor's degree)** | | | |
| | High school diploma or less | 3.43 (2.11, 5.59) | <.001 |
| | College or some college | 1.87 (1.19, 2.95) | .001 |
| **Caregiver status (reference: Not a caregiver)** | | | |
| | Caregiver for children only | .91 (.60, 1.39) | >.99 |
| | Caregiver for adults only | .75 (.52, 1.09) | .32 |
| | Caregiver for adults & children | .51 (.35, .74) | <.001 |
| **Political ideology (reference: Very liberal)** | | | |
| | Slightly liberal | .92 (.54, 1.56) | >.99 |
| | Neither liberal nor conservative | 2.74 (1.73, 4.32) | <.001 |
| | Slightly conservative | 1.99 (1.23, 3.23) | .001 |
| | Very conservative | 3.58 (2.16, 5.94) | <.001 |
| | Apolitical or prefer not to say | 3.27 (1.86, 5.75) | <.001 |
| **Mask-wearing in public (reference: Always or Often)** | | | |
| | Sometimes | 2.34 (1.50, 3.64) | <.001 |
| | Rarely or Never | 3.92 (2.52, 6.10) | <.001 |
| **Avoid gatherings (reference: Always or Often)** | | | |
| | Sometimes | 1.29 (.86, 1.93) | .85 |
| | Rarely or Never | 2.65 (1.95, 3.60) | <.001 |
| **Medical Mistrust Index (reference: 0-6)** | | | |
| | 7-13 | .88 (.51, 1.52) | >.99 |
| | 14-17 | 1.41 (.81, 2.43) | .85 |
| | 18-21 | 2.11 (1.10, 4.07) | .014 |
| **Received or plan to receive influenza vaccine this year (reference: Yes)** | | | |
| | No or not sure | 4.11 (3.05, 5.54) | <.001 |

Adjusted odds ratios (aORs) and 95% confidence intervals (CIs) are shown in Figure 1.

### eTable 2. Unadjusted odds ratios for COVID-19 vaccine refusal among US adults—March 2021

| | | Vaccine Refusal | |
|---|---|---|---|
| | | **OR (95% CI)** | ***P*** |
| **Sex (reference: Male)** | | | |
| | Female | 1.68 (1.32, 2.15) | <.001 |
| **Age group, years (reference: ≥65)** | | | |
| | 18-24 | 3.37 (1.81, 6.29) | <.001 |
| | 25-44 | 2.70 (1.59, 4.59) | <.001 |
| | 45-64 | 2.60 (1.51, 4.48) | <.001 |
| **Pregnancy (reference: non-pregnant females aged 18-49 years)** | | | |
| | Yes | .44 (.28, .69) | <.001 |
| **Race/ethnicity (reference: White, non-Hispanic)** | | | |
| | Black, non-Hispanic | 1.40 (1.02, 1.93) | .031 |
| | Asian, non-Hispanic | .41 (.21, .80) | .002 |
| | Other, non-Hispanic | 1.92 (1.17, 3.13) | .002 |
| | Hispanic, any race or races | .83 (.52, 1.34) | >.99 |
| **Education (reference: More than Bachelor's degree)** | | | |
| | High school diploma or less | 5.61 (3.56, 8.84) | <.001 |
| | College or some college | 2.35 (1.52, 3.64) | <.001 |
| **Caregiver status (reference: Not a caregiver)** | | | |
| | Caregiver for children only | 1.12 (.76, 1.64) | >.99 |
| | Caregiver for adults only | .83 (.59, 1.16) | >.99 |
| | Caregiver for adults & children | .51 (.37, .71) | <.001 |
| **Political ideology (reference: Very liberal)** | | | |
| | Slightly liberal | .87 (.53, 1.41) | >.99 |
| | Neither liberal nor conservative | 2.72 (1.79, 4.12) | <.001 |
| | Slightly conservative | 1.54 (.99, 2.41) | .063 |
| | Very conservative | 2.66 (1.69, 4.19) | <.001 |
| | Apolitical or prefer not to say | 4.93 (2.83, 8.57) | <.001 |
| **Mask-wearing in public (reference: Always or Often)** | | | |
| | Sometimes | 2.18 (1.49, 3.17) | <.001 |
| | Rarely or Never | 4.04 (2.72, 6.01) | <.001 |
| **Avoid gatherings (reference: Always or Often)** | | | |
| | Sometimes | 1.23 (.86, 1.77) | >.99 |
| | Rarely or Never | 2.99 (2.27, 3.94) | <.001 |
| **Medical Mistrust Index (reference: 0-6)** | | | |
| | 7-13 | .90 (.54, 1.48) | >.99 |
| | 14-17 | 1.35 (0.80, 2.26) | >.99 |
| | 18-21 | 1.86 (1.01, 3.43) | .045 |
| **Received or plan to receive influenza vaccine this year (reference: Yes)** | | | |
| | No or not sure | 5.86 (4.39, 7.81) | <.001 |

### eTable 3. Adjusted odds ratios for responding Maybe to obtaining a COVID-19 vaccine as soon as possible among US adult vaccine Refusers—March 2021

| | | Maybe | |
|---|---|---|---|
| | | **aOR (95% CI)** | ***P* value** |
| **Sex (reference: Male)** | | | |
| | Female | .89 (.63, 1.26) | >.99 |
| **Age group, years (reference: ≥65)** | | | |
| | 18-24 | 2.30 (1.08, 4.90) | .021 |
| | 25-44 | 1.41 (.75, 2.64) | >.99 |
| | 45-64 | 1.43 (.76, 2.69) | .87 |
| **Pregnancy (reference: non-pregnant females aged 18-49 years)** | | | |
| | Yes | 1.88 (.92, 3.87) | .13 |
| **Race/ethnicity (reference: White, non-Hispanic)** | | | |
| | Black, non-Hispanic | 1.11 (.69, 1.79) | >.99 |
| | Asian, non-Hispanic | 2.63 (.93, 7.42) | .086 |
| | Other, non-Hispanic | .95 (.46, 1.98) | >.99 |
| | Hispanic, any race(s) | 1.08 (.62, 1.90) | >.99 |
| **Education (reference: More than Bachelor's degree)** | | | |
| | High school diploma or less | .42 (.22, .81) | .003 |
| | College or some college | .68 (.36, 1.27) | .69 |
| **Caregiver status (reference: Not a caregiver)** | | | |
| | Caregiver for children only | .96 (.57, 1.63) | >.99 |
| | Caregiver for adults only | 1.03 (.63, 1.69) | >.99 |
| | Caregiver for adults & children | 1.58 (1.01, 2.47) | .042 |
| **Political ideology (reference: Very conservative)** | | | |
| | Slightly conservative | 2.17 (1.27, 3.69) | .001 |
| | Center | 2.57 (1.61, 4.12) | <.001 |
| | Slightly liberal | 2.92 (1.58, 5.42) | <.001 |
| | Very liberal | 1.39 (.69, 2.80) | >.99 |
| | Apolitical or prefer not to say | 2.13 (1.13, 4.03) | .009 |

**eTable 4. Unadjusted odds ratios for responding Maybe to obtaining a COVID-19 vaccine as soon as possible among US adult vaccine Refusers —March 2021**

| | | Maybe | |
|---|---|---|---|
| | | **OR (95% CI)** | ***P* value** |
| **Sex (reference: Male)** | | | |
| | Female | .81 (.57, 1.13) | .62 |
| **Age group, years (reference: ≥65)** | | | |
| | 18-24 | 2.87 (1.38, 5.99) | .001 |
| | 25-44 | 1.68 (.91, 3.08) | .16 |
| | 45-64 | 1.60 (.85, 3.00) | .33 |
| **Pregnancy (reference: non-pregnant females aged 18-49 years)** | | | |
| | Yes | 2.79 (1.49, 5.24) | <.001 |
| **Race/ethnicity (reference: White, non-Hispanic)** | | | |
| | Black, non-Hispanic | 1.29 (.81, 2.06) | .94 |
| | Asian, non-Hispanic | 3.12 (1.23, 7.95) | .007 |
| | Other, non-Hispanic | 1.05 (.53, 2.07) | >.99 |
| | Hispanic, any race(s) | 1.24 (.67, 2.28) | >.99 |
| **Education (reference: More than Bachelor's degree)** | | | |
| | High school diploma or less | .40 (.21, .75) | .001 |
| | College or some college | .60 (.32, 1.11) | .18 |
| **Caregiver status (reference: Not a caregiver)** | | | |
| | Caregiver for children only | 1.04 (.61, 1.75) | >.99 |
| | Caregiver for adults only | 1.14 (.72, 1.79) | >.99 |
| | Caregiver for adults & children | 1.97 (1.28, 3.05) | <.001 |
| **Political ideology (reference: Very conservative)** | | | |
| | Slightly conservative | 2.33 (1.37, 3.98) | <.001 |
| | Center | 2.72 (1.70, 4.36) | <.001 |
| | Slightly liberal | 3.23 (1.76, 5.93) | <.001 |
| | Very liberal | 1.59 (.82, 3.08) | .41 |
| | Apolitical or prefer not to say | 1.93 (.99, 3.79) | .058 |

**eTable 5. Adjusted odds ratios for waiting for more safety and efficacy data before obtaining a COVID-19 vaccine among US adult vaccine Refusers —March 2021**

| | | Maybe | |
|---|---|---|---|
| | | aOR (95% CI) | *P* value |
| **Sex (reference: Male)** | | | |
| | Female | 1.04 (.71, 1.52) | >.99 |
| **Age group, years (reference: ≥65)** | | | |
| | 18-24 | .39 (.15, 1.00) | .051 |
| | 25-44 | .41 (.19, .89) | .013 |
| | 45-64 | .49 (.24, 1.01) | .06 |
| **Pregnancy (reference: non-pregnant females aged 18-49 years)** | | | |
| | Yes | 1.10 (.50, 2.44) | >.99 |
| **Race/ethnicity (reference: White, non-Hispanic)** | | | |
| | Black, non-Hispanic | .83 (.47, 1.44) | >.99 |
| | Asian, non-Hispanic | 2.66 (.97, 7.25) | .06 |
| | Other, non-Hispanic | .48 (.20, 1.15) | .16 |
| | Hispanic, any race(s) | .93 (.44, 1.95) | >.99 |
| **Education (reference: More than Bachelor's degree)** | | | |
| | High school diploma or less | .53 (.26, 1.09) | .12 |
| | College or some college | .99 (.50, 1.97) | >.99 |
| **Caregiver status (reference: Not a caregiver)** | | | |
| | Caregiver for children only | 1.23 (.71, 2.12) | >.99 |
| | Caregiver for adults only | 1.23 (.74, 2.06) | >.99 |
| | Caregiver for adults & children | 1.39 (.82, 2.35) | .63 |
| **Political ideology (reference: Very conservative)** | | | |
| | Slightly conservative | 1.49 (.83, 2.67) | .48 |
| | Center | 1.08 (.63, 1.83) | >.99 |
| | Slightly liberal | 1.60 (.81, 3.16) | .45 |
| | Very liberal | .88 (.42, 1.83) | >.99 |
| | Apolitical or prefer not to say | .98 (.45, 2.14) | >.99 |

### eTable 6. Unadjusted odds ratios for waiting for more safety and efficacy data before obtaining a COVID-19 vaccine among US adult vaccine Refusers —March 2021

| | | Maybe | |
|---|---|---|---|
| | | **OR (95% CI)** | ***P* value** |
| **Sex (reference: Male)** | | | |
| | Female | .94 (.63, 1.40) | >.99 |
| **Age group, years (reference: ≥65)** | | | |
| | 18-24 | .45 (.19, 1.07) | .092 |
| | 25-44 | .44 (.20, .95) | .031 |
| | 45-64 | .52 (.24, 1.15) | .19 |
| **Pregnancy (reference: non-pregnant females aged 18-49 years)** | | | |
| | Yes | 1.34 (.68, 2.66) | >.99 |
| **Race/ethnicity (reference: White, non-Hispanic)** | | | |
| | Black, non-Hispanic | .83 (.48, 1.45) | >.99 |
| | Asian, non-Hispanic | 2.63 (1.10, 6.31) | .021 |
| | Other, non-Hispanic | .56 (.24, 1.30) | .44 |
| | Hispanic, any race(s) | 1.06 (.47, 2.41) | >.99 |
| **Education (reference: More than Bachelor's degree)** | | | |
| | High school diploma or less | .52 (.25, 1.08) | .11 |
| | College or some college | 1.00 (.51, 1.94) | >.99 |
| **Caregiver status (reference: Not a caregiver)** | | | |
| | Caregiver for children only | 1.07 (.61, 1.87) | >.99 |
| | Caregiver for adults only | 1.20 (.71, 2.02) | >.99 |
| | Caregiver for adults & children | 1.18 (.73, 1.93) | >.99 |
| **Political ideology (reference: Very conservative)** | | | |
| | Slightly conservative | 1.38 (.79, 2.41) | .84 |
| | Center | .94 (.58, 1.51) | >.99 |
| | Slightly liberal | 1.37 (.72, 2.60) | >.99 |
| | Very liberal | .74 (.38, 1.44) | >.99 |
| | Apolitical or prefer not to say | .80 (.29, 2.23) | >.99 |
